# Feasibility of Monkeypox virus sequencing from antigen rapid diagnostic tests as a potential tool to enhance genomic surveillance

**DOI:** 10.64898/2026.07.09.26356424

**Authors:** Charlotte Pronier, Adriana Renzoni, Florian Laubscher, Valentin Chudzinski, Kenneth Adea, Placide Mbala-Kingebeni, Camille Escadafal, Isabella Eckerle

**Affiliations:** Laboratory of Virology, CHU Rennes, Rennes, France; Univ Rennes, Inserm, EHESP, Irset (Institut de recherche en santé, environnement et travail) - UMR_S 1085, F-35000 Rennes, France; Laboratory of Virology, Laboratory Medicine Division, Diagnostic Department, Geneva University Hospitals; Geneva, Switzerland; Geneva Center for Emerging Viral Diseases, Geneva University Hospitals and Faculty of Medicine, University of Geneva, Geneva, Switzerland; Department of Medicine, Faculty of Medicine, University of Geneva, Geneva, Switzerland; Department of Microbiology and Molecular Medicine, Faculty of Medicine, University of Geneva, Geneva, Switzerland; University of Kinshasa Medical School, Democratic Republic of the Congo; Institut National de Recherche Biomédicale, Democratic Republic of the Congo; South African National Bioinformatics Institute, University of the Western Cape, Republic of South Africa; Department of Virology, Graduate School of Medicine, Osaka Metropolitan University, Japan

**Keywords:** Monkeypox virus, mpox, sequencing, metagenomics, rapid diagnostic tests

## Abstract

**Background:** Sequencing of monkeypox virus (MPXV) from antigen rapid diagnostic tests (Ag-RDTs) could expand genomic surveillance during outbreaks in decentralized settings where sequencing equipment and cold chain transportation are unavailable. We aimed to evaluate the efficacy of MPXV sequencing from MPXV antigen Ag-RDTs.

**Methods:** We tested MPXV Ag-RDTs from three different brands using serial dilutions of cultured MPXV subclade Ib. Positive Ag-RDTs with different intensities of the test band were stored for 19 days, either at room temperature or at +4°C, after which viral DNA was extracted from the pads of the test cassettes. Metagenomic and tiled amplicon-based Oxford Nanopore technology sequencing methods were then performed.

**Results:** Viral DNA extraction from MPXV Ag-RDTs showed a consistent decrease in viral load of 3 logs compared to the initial viral load of the applied viral dilution. Both sequencing methods were able to reach high coverage but the tiled amplicon-based demonstrated more consistent results with a coverage always above 85%.

**Conclusion:** This proof-of-concept supports the development of this approach in the field, with the aim of combining genomic surveillance with decentralized testing, including in remote areas.

**Importance statement:** As monkeypox virus (MPXV) continues to expand its circulation, genomic surveillance of MPXV is essential to monitor viral evolution, detect emerging variants, guide control measures and adapt public health responses. However, diagnostic and sequencing capacity is limited in many regions where mpox is endemic, and transporting clinical samples from remote testing sites to reference laboratories remains logistically challenging. Rapid antigen diagnostic tests (Ag-RDTs) now have the potential to be increasingly used for decentralized mpox testing and could represent an alternative source of viral genetic material for sequencing. In this study, we demonstrate that MPXV DNA can be recovered from used Ag-RDT devices and successfully sequenced using Oxford Nanopore technologies, even after 19 days of storage at room temperature. Importantly, in almost all tested conditions, genome coverage was sufficient for viral subclade identification and mutation analysis. These findings highlight the potential of Ag-RDTs not only as diagnostic tools but also as practical sampling devices for genomic surveillance, which could significantly expand sequencing capacity in resource-limited and remote settings.

## Introduction

Mpox is a viral zoonotic and emerging disease caused by the monkeypox virus (MPXV), a species of the genus *Orthopoxvirus* within the *Poxviridae* family. MPXV possesses a linear double-stranded DNA genome of approximately 200 kilobase pairs encoding around 200 proteins, making it one of the largest known viral genomes. The central region of the genome is highly conserved and contains essential replication genes, while the terminal regions are more variable and enriched in genes associated with host interaction and pathogenicity. Based on phylogenetic analysis, MPXV is divided into two clades, which are themselves subdivided into subclades: clade I, with subclades Ia and Ib and clade II, with subclades IIa and IIb.

Historically, MPXV has been characterized as an endemic zoonotic disease transmitted through contact with rodent reservoirs. However, 91’000 confirmed cases were reported in 116 non-endemic countries during a global epidemic of MPXV subclade IIb in 2022(1),(2),(3), demonstrating spread of the disease beyond countries with known reservoirs. MPXV sequences from 2022 revealed a series of mutations characteristic of host enzymes with antiviral function called APOBEC3 deaminases. A study considering APOBEC3 editing as a genetic marker of human infection, estimates that MPXV has been circulating in humans since 2016(4). In late 2023, an increase in cases of a novel subclade, named Ib, was observed in the Democratic Republic of the Congo (DRC) and subsequently in other African countries. This subclade displays new mutations in the C9L and B21R genes, that enable the virus to adapt more effectively to human hosts (5). The emergence of these MPXV clade I viruses enabling human transmission, and the potential of these viruses to recombine between themselves, as observed in India and the UK in 2025 (6, 7), argue for a strengthened global genetic surveillance of circulating MPXV. Although Africa has lifted MPXV as a public health emergency of continental security (8), all clades of MPXV continue to circulate and there is a risk of sustained community transmission. In 2025, there were 52’845 confirmed MPXV cases with 215 deaths reported from 98 countries worldwide.

Laboratory confirmation is crucial for precise case identification, timely patient management, and informed public health responses. The gold-standard method for detecting MPXV is PCR, which provides high accuracy and specificity. However, PCR testing requires complex laboratory infrastructure and trained personnel, making it largely confined to centralized laboratory settings (9). Some molecular point-of-care assays have been developed for MPXV; however, these remain expensive and/or lack independent performance data. Currently, no CE-IVD marked nucleic acid amplification test allows for the rapid diagnosis of MPXV and the determination of all subclades (Ia, Ib, IIa, and IIb). Therefore, genome sequencing remains the main technique to determine the subclade responsible for an mpox infection or outbreak, and to monitor viral evolution.

Given these limitations, there is an urgent need to expand and decentralize MPXV testing to improve its accessibility, especially in low- and middle-income countries (LMICs). Rapid antigen-detection tests (Ag-RDTs) represent a promising approach that could address this gap. Ag-RDTs are easy to use, provide quick results, require minimal equipment, and can be deployed at the point-of-care and at the community level.

As per the FIND test directory, 32 Ag-RDTs are commercially available and have received regulatory approval(10). However, clinical performance data is scarce and the reported sensitivity values do not meet the minimum criteria of the target product profile (TPP) for tests used as aid to mpox diagnosis published by WHO (11–13). Global efforts are made to improve the sensitivity of Ag-RDTs and related data, as there is a significant potential in integrating them into mpox diagnostic strategies, especially in resource-limited environments, complementing PCR testing and supporting more effective outbreak control.

Genomic surveillance of monkeypox virus was initiated in 2022 due to the unprecedented human-to-human spread of MPXV. This surveillance is crucial for guiding public health interventions, given that MPXV clades have demonstrated differences in virulence and case fatality rates, and that certain variants may be resistant to antiviral drugs such as tecovirimat (14, 15). Genomic surveillance is also an essential tool for epidemiological investigations and for identifying known or novel mutations. However, sequencing-based surveillance is very limited in low-resource or remote areas, since it requires expensive equipment, maintenance, and trained personnel as well as efficient sample transportation. One approach that could help to overcome these limitations would be to use Ag-RDT strips collected at decentralized sites and sent to centralized reference laboratories for genomic analysis, rather than collecting, storing, and shipping an additional swab sample.

With the inclusion of lateral-flow assays and alternative sampling matrices (e.g. dried blood spots, used paper tissues, etc…) in diagnostic strategies, there is an opportunity to consider these materials for genomic surveillance(16, 17). These alternative sample types offer practical advantages as they are easier to collect, store, and transport than traditional clinical samples such as blood or swabs in a transport medium. They often require less stringent cold chain management and can remain stable at ambient temperatures for extended periods. Overall, they expand the reach and feasibility of viral infection monitoring beyond conventional clinical settings. As an example, dried blood spots are widely used for molecular surveillance of viruses such as HIV and hepatitis viruses. To our knowledge, whole genome sequencing from Ag-RDT strips has already been performed for SARS-CoV-2 and other respiratory viruses (influenza, RSV) or dengue virus, but not for MPXV(18–22).

In this context, we performed a proof-of-concept study to evaluate the reliability of Ag-RDTs in generating MPXV sequencing data by assessing two types of sequencing protocols, three brands of mpox Ag-RDTs and two storage temperature conditions.

## Materials and Methods

### Virus propagation

A MPXV subclade Ib isolate was shared by the Spiez Laboratory in Switzerland via the WHO BioHub System (reference number 2024-WHO-LS-003) and originated from a patient in DRC. The MPXV subclade Ib was propagated on Vero-E6 cells at the University Hospitals of Geneva under Biosafety Level 3 conditions, as described previously(13). The resulting cultured virus was sequenced and the sequence uploaded to Genbank under accession number PX049155.

### MPXV Ag-RDT testing

The three Ag-RDT brands used in this study were: NG-Test® Monkeypox virus (NG Biotech, Guipry, France), Monkey Pox Antigen Test Cassette (Testsealabs®, Hangzhou, China) and Monkeypox Virus Antigen Rapid Test Kit (Contipharma, Seraing, Belgium). The main characteristics of these tests are presented in Table S1. We performed tests in the BSL3 laboratory and according to the manufacturer’s instructions with the exception that 200 μL of virus dilution was added directly to the buffer tube instead of a swab specimen.

For each Ag-RDT brand, we tested 3 viral dilutions with Ct values of 23.1 (1.21E+07 copies/ml), 24.5 (3.81E+06 copies/ml) and 26.4 (1.31E+06 copies/ml). After Ag-RDT loading on the sample pad, we observed 3 different band intensities, which were defined as high, medium and low intensities. For each test brand and each band intensity, 2 storage temperatures were evaluated: room temperature around 21°C (RT) and refrigerated (+4°C).

After 19 days at RT or +4°C, each Ag-RDT cassette was opened, and the sample and conjugation pads were extracted (Figure 1). For each Ag-RDT, both pads were placed in a 1.5 ml Eppendorf tube containing 500 µl of NUCLISENS® easyMAG® Lysis Buffer (bioMérieux), incubated 10 minutes at RT followed by a 10 min incubation at 70°C. For the NG-Biotech RDTs, this treatment resulted in a highly viscous solution. We therefore performed DNA extraction by adding 500 µl of PBS to the pad, mixed gently for 30 minutes and the resulting supernatant was added to 500 µl of the lysis buffer. After centrifugation of the lysed solution (1000 µl for NG-Biotech RDT; 500 µl for Contipharma and Testsea RDTs) at 10,000 rpm for 5min, DNA was extracted from the inactivated samples using the eMAG automated extractor from bioMérieux, according to the manufacturer’s instructions, with an elution volume of 25 µl.

**Figure 1.**
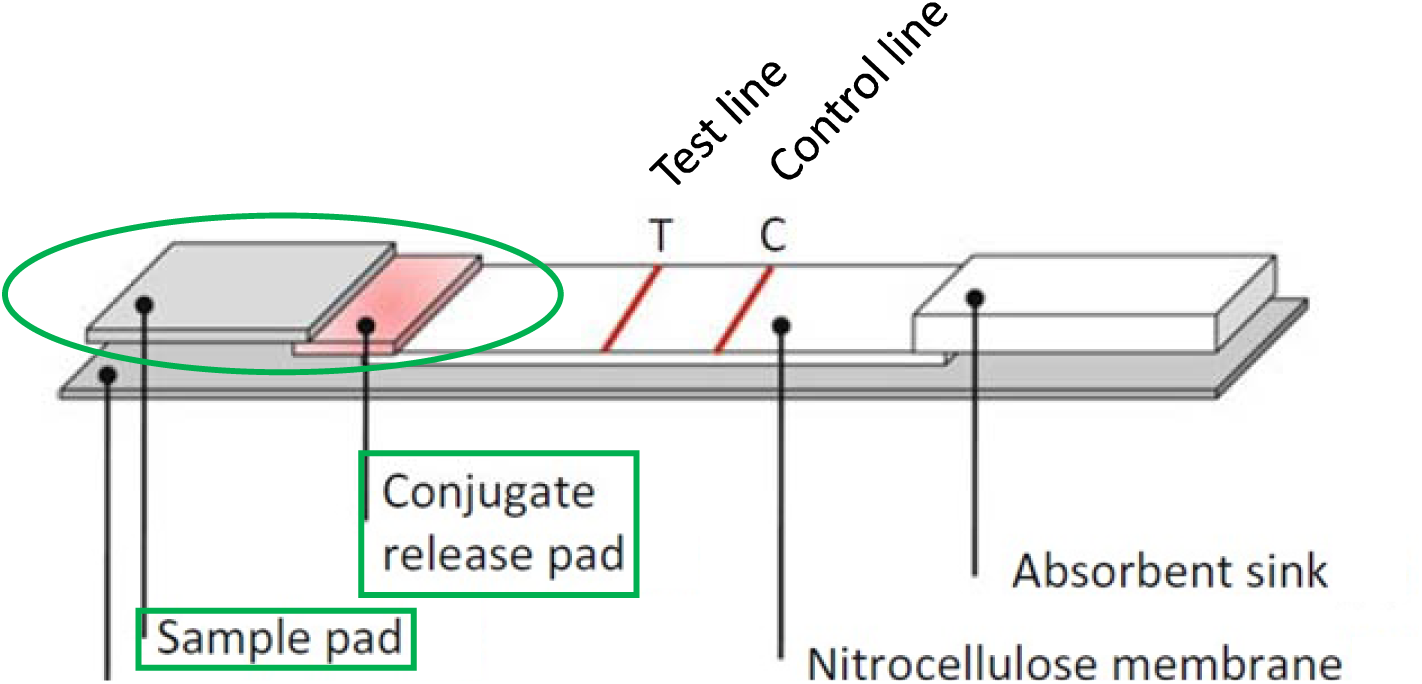
Structural overview of the components inside a rapid test device cassette. The sample and conjugation pads were both used for nucleic acid extraction (circled in green).

An in-house real-time *Orthopoxvirus* PCR was used to obtain cycle threshold values for each dilution and after strip elution of each Ag-RDT(23)(24).

### Sequencing

We tested two different approaches to sequence MPXV whole genome in parallel from the same nucleic acid extract: one metagenomic-based and one tiled amplicon-based technique.

For metagenomic-based method, sequencing libraries were generated using the rapid sequencing DNA PCR Barcoding Kit (SQK-RPB114.24, Oxford Nanopore Technologies) according to the manufacturer’s instructions. We used an input DNA volume of 3 µl and increased the number of amplification cycles to 25, before sequencing on a MinION flow cell (FLO-MIN114 R10, Oxford Nanopore Technologies) for at least 50 hours.

For the amplicon-based method, we used the protocol designed by Welkers et al., which consists of a PCR-based amplicon tiling approach involving a total of 88 primer sets divided between 2 amplicon pools(25). The amplicon size is approximately 2.5kB. We used the NextGenPCR^TM^ MPXV sequencing library prep kit 96RXN (RUO, Molecular Biology Systems, the Netherlands), composed of Arctic Fox HF Chemistry-2X, a ready-to-use polymerase master mix, and the 2 MPXV premixed primer pools. PCR was performed with the Biometra thermocycler. Amplicon-based libraries were generated using the rapid sequencing DNA V14 kit (SQK-RBK114.24, Oxford Nanopore Technologies), according to the manufacturer’s recommendations, before sequencing on a MinION flow cell (FLO-MIN114 R10, Oxford Nanopore Technologies) for at least 46 hours. A negative control was included and processed in parallel with samples to monitor potential contamination.

### Bioinformatics

Raw sequencing reads in FASTQ format were first processed to remove any remaining adapter sequences using Porechop. In cases where the sequencing data originated from amplicon-based protocols, the pipeline was executed in amplicon-aware mode (enabled with the -a flag). This mode uses a modified version of Porechop (https://github.com/Laubscher/porexop) to trim reads based on provided primer sequences.

Trimmed reads were then aligned to a database of genomes representing the four MPXV subclades using minimap2. For each sample, the best-matching genome was automatically selected based on the total consensus length. Final consensus genome sequences were generated using samtools and iVar, retaining only regions with a minimum coverage depth of 10× and filtering insertions with a minimum frequency threshold of at least 50% of the coverage at a given position. Positions with insufficient coverage were masked with ambiguous bases (N). All scripts used in this pipeline are openly accessible at https://github.com/Laubscher/MPXV. The overwiew of the bioinformatics pipeline used for MPXV sequencing is mapped in Figure S1. The resulting consensus FASTA sequences were then analyzed with Nextclade (https://clades.nextstrain.org/) using the nextstrain/mpox/all-clade dataset to assign clades.

Sample sequences were aligned and compared against the reference sequence derived from the isolate (GenBank accession number PX049155) (Figure S2).

## Results

### Efficiency of DNA extraction from sample pads and correlation with band intensities.

In total, we tested 18 Ag-RDT strips, each of which corresponded to a different set of conditions relating to the 3 dilution concentrations, the 3 Ag-RDT brands and the 2 storage temperatures. Each Ag-RDT strip was tested with the 2 sequencing methods but from the same nucleic acid extract.

To assess the efficiency of DNA extraction from pads of different Ag-RDT brands, we compared Ct values of each viral dilution before running on the pad (initial Ct values) and after pad elution (Table S2). The initial Ct values and viral loads for the three tested dilutions corresponding to high-, medium- and low-intensity bands were 23.1 (quantified as 1.21E+07 copies/ml or 7.1 log copies/ml), 24.8 (quantified as 3.81E+06 copies/ml or 6.6 log copies/ml), and 26.4 (quantified as 1.31E+06 copies/ml or 6.1 log copies/ml), respectively. Following Ag-RDT strip elution, viral DNA was amplified, yielding Ct values ranging from 26.3 to 27.8 for high-intensity bands, 28.1 to 31.5 for medium-intensity bands and 30.4 to 32 for low-intensity bands (Figure 2). A significant correlation was observed between the initial dilution Ct values and those obtained after strip elution (R^2^=0.897, p < 0.0001), with no effect of Ag-RDT storage temperature.

**Figure 2.**
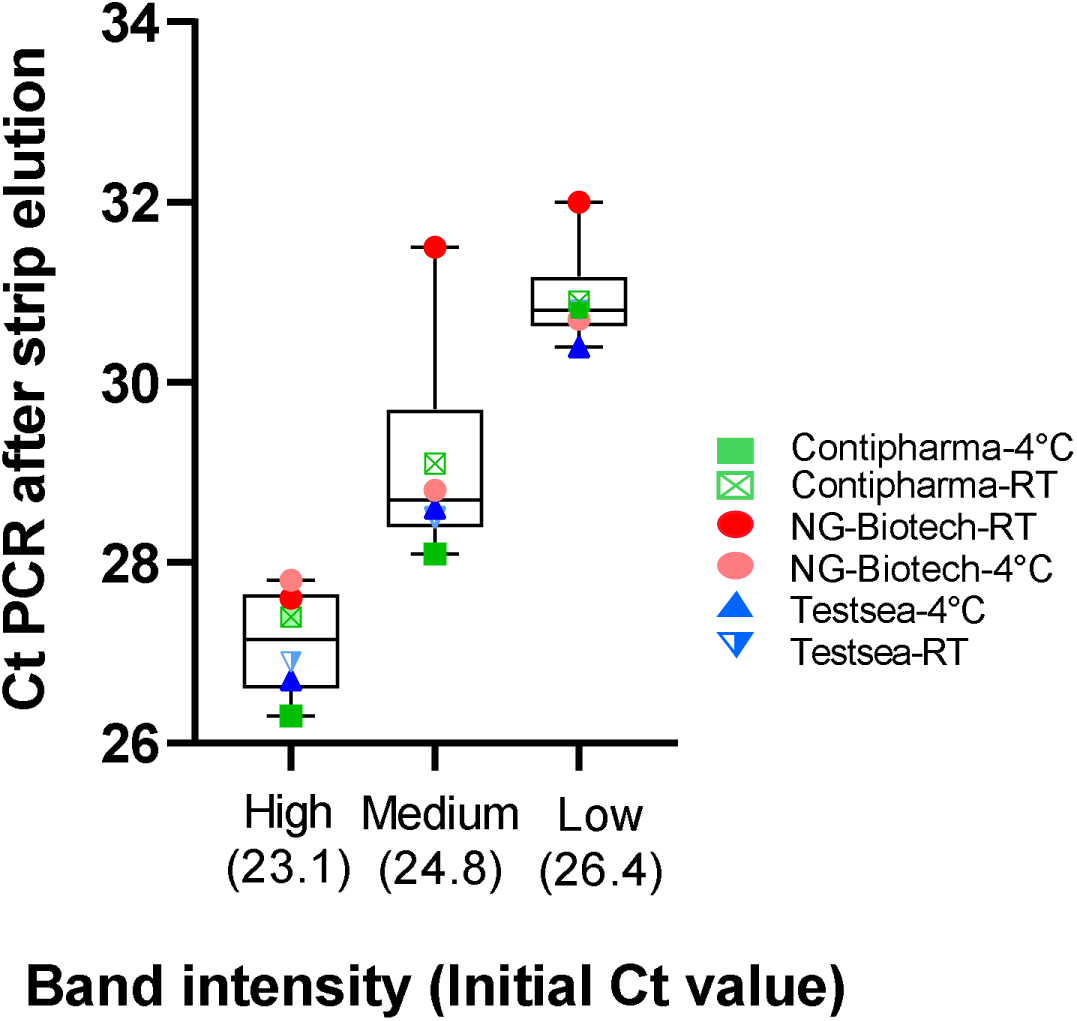
Cycle threshold (Ct) values after strip elution according to the level of band intensity and initial threshold values.

### Genome Mpox sequencing by metagenomic and tiling methods after pad DNA extraction.

The same nucleic acid extract from each pad was sequenced by the 2 different sequencing methods. MPXV sequences were successfully obtained and allowed to identify the subclade as Ib in all but one of the 36 tested conditions (Table 1). However, analyzing sequencing depth and genome coverage, some differences were observed (Table 1; Figure 3). The median sequencing depth was higher with the tiling method than with the metagenomics method. For the Ag-RDTs stored at RT, the median depth was 1009 (range 785) versus 160 (range 1543) using the tiling and metagenomic method, respectively. For the Ag-RDTs stored at +4°C, the median depths were 961 (range 1075) and 35 (range 830), respectively. Notably, the tiling method achieved genome coverage above 85% in all testing conditions and coverage exceeded 90% in 14 out of 18 tested conditions (77.8%). In contrast, the metagenomics method achieved >70% coverage in 9 out of 18 tested conditions (50%).

**Figure 3.**
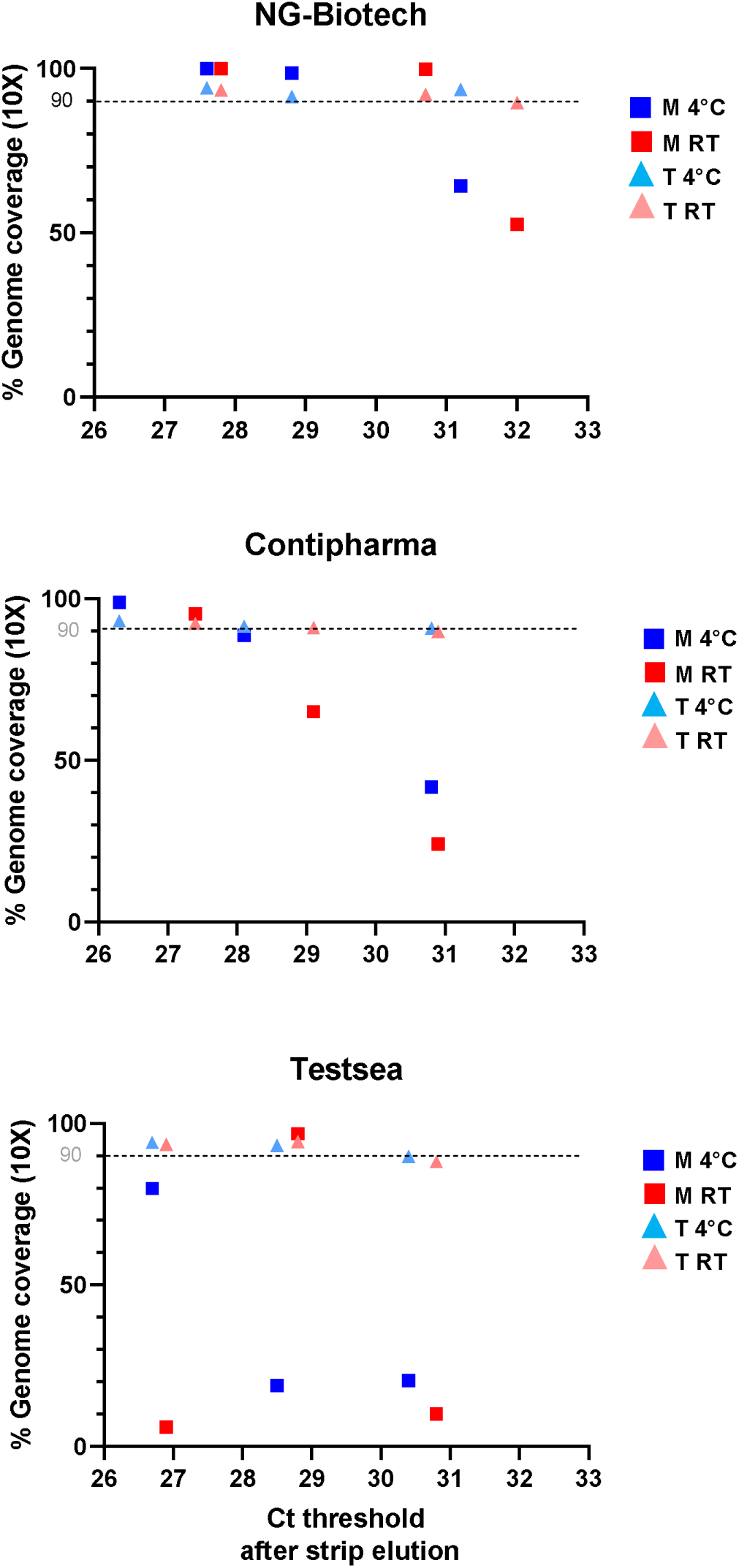
Comparison of percent genome coverage at 10X of specimen sequenced with metagenomic (M) and tiling (T)-based methods according to Ct value measured by PCR after RDT pad elution and storage temperature, either 4°C or room temperature (RT) for Contipharma, NG-Biotech and Testsea RDTs.

**Table 1.** Capacity to identify the MPXV subclade, median depth and percent of genome coverage at 10x according to the sequencing method, the storage temperature (RT/+4°C), the intensity of the test band (high/medium/low) and the Ag-RDT brand (Contipharma/NG Biotech/Testsea).

| Band intensity | Ag-RDT brand | Subclade identification |  |  |  | Median depth |  |  |  | Genome coverage % (10x) |  |  |  |
| --- | --- | --- | --- | --- | --- | --- | --- | --- | --- | --- | --- | --- | --- |
|  |  | RT-Meta | RT-Tiling | 4°C-Meta | 4°C-Tiling | RT-Meta | RT-Tiling | 4°C-Meta | 4°C-Tiling | RT-Meta | RT-Tiling | 4°C-Meta | 4°C-Tiling |
| High | NG-Biotech | yes | yes | yes | yes | 831 | 1 414 | 1 572 | 1 318 | 99.9% | 94.1% | 99.9% | 93.4% |
| High | Contipharma | yes | yes | yes | yes | 185 | 887 | 160 | 1 455 | 95.3% | 92.4% | 98.9% | 93.2% |
| High | Testsea | no | yes | yes | yes | 1 | 949 | 87 | 1 000 | 1.6% | 93.5% | 79.9% | 94.1% |
| Medium | NG-Biotech | yes | yes | yes | yes | 90 | 1 726 | 663 | 905 | 64.3% | 93.5% | 98.5% | 91.4% |
| Medium | Contipharma | yes | yes | yes | yes | 35 | 961 | 78 | 768 | 65.0% | 91.0% | 88.7% | 91.5% |
| Medium | Testsea | yes | yes | yes | yes | 62 | 1 272 | 195 | 1 553 | 18.9% | 93.2% | 96.9% | 94.4% |
| Low | NG-Biotech | yes | yes | yes | yes | 34 | 991 | 254 | 1 524 | 52.5% | 89.6% | 99.7% | 92.0% |
| Low | Contipharma | yes | yes | yes | yes | 32 | 789 | 35 | 1 009 | 24.1% | 89.9% | 41.7% | 90.9% |
| Low | Testsea | yes | yes | yes | yes | 1 | 651 | 29 | 848 | 5.1% | 88.2% | 20.4% | 89.8% |

To determine whether storage temperature influences sequencing results, we analyzed genome coverage for both sequencing methods. With the tiling method, coverage remained constant across temperatures, with no significant difference in median genome coverage between RT [92.4% with SD:3.7%) and refrigerated storage (92% with SD:2.4%) (p = 0.2153). In contrast, genome coverage obtained with the metagenomic method differed significantly. Coverage was lower in RT-stored pads (52.5% with SD:60.8%) compared to those with refrigerated storage (96.9% with SD:32%) (p = 0.0095).

When the genome coverage percentages obtained with both sequencing methods are compared across the three Ag-RDTs, the Contipharma Ag-RDT results demonstrate a coverage percentage of over 65% with either sequencing method for all samples with high and medium band intensities, while the coverage percentage was lower for the 2 samples with low band intensity (Ct values > 30 after pad elution) (Figure 3). With the NG Biotech Ag-RDTs, the genome coverage is greater than 50% for all testing conditions. With the Testsea Ag-RDTs, the differences in genome coverage according to the sequencing approach are greater. The genome coverage was greater than 85% for all 6 samples with the tiling method and for only one sample with the metagenomic method. Good quality genome coverage was obtained with all three Ag-RDTs brands tested.

### In depth analysis of metagenomic and tiling methods

Comparison of the two sequencing approaches revealed distinct coverage patterns across the MPXV genome (Figure 4). Although the tiled-amplicon method produced largely uniform coverage, recurrent dropouts were consistently observed for six amplicons. For most of them, the loss of coverage corresponded to one or two mismatches in either the left or right primer. In two other cases (amplicons 59 and 70), the dropout resulted from a specific genomic deletion present in the Ib MPXV strain, spanning the primer-binding region. In contrast, the metagenomic approach showed more variable coverage within the central genomic regions but achieved higher depth and more complete recovery at the genome extremities. It is interesting to note that in our testing conditions, coverage as low as 5.1% was sufficient to determine the MPXV subclade.

**Figure 4.**
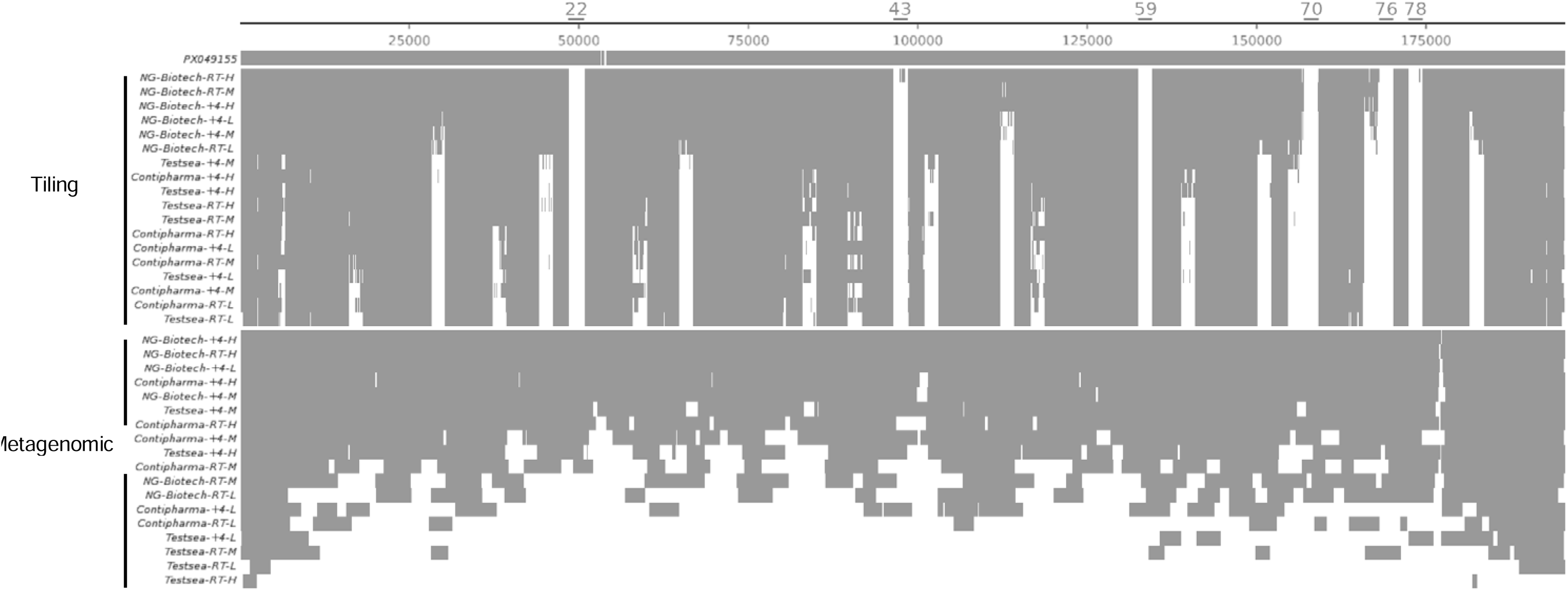
Genome coverage overview and gaps for each of the 36 tested conditions compared to the reference genome CMVE-2 (PX049155). Amplicons with defective amplification are labeled by their corresponding numbers above.

### Difference with the reference sequence

Overall, the obtained sequences are highly similar to the reference sequence, with only a few differences observed. We observed variability in the number of TA repeat motifs around position ∼177,120 bp. The number of repeats varied substantially across the tested conditions, reaching up to 41 TA repeats in our dataset. This pattern was exclusively detected with the tiled-amplicon approach; sequences obtained with the metagenomic approach showed little to no coverage in this genomic region, preventing reliable characterization of the number of repeats. For comparison, the reference sequence PX049155 contains 6 TA repeats, while the sequence (GISAID ID: EPI_ISL_19348512) of the first mpox case with MPXV clade Ib outside Africa reported in Sweden harbors 59 repeats(26).

Across the 36 tested conditions, the tiling-amplicon sequencing approach produced consensus genomes identical to the reference sequence (PX049155), with zero single-nucleotide polymorphisms (SNP) differences when excluding uncovered (N) regions, ambiguous positions, and the TA repeat region (18/18 conditions). In contrast, metagenomic sequencing yielded zero SNPs for only 6 of 18 conditions, while the remaining 12 conditions showed 1 to 8 SNPs relative to the reference sequence under the same criteria (supplemental figure S1). These differences remained very small, and it is not possible to determine whether the observed SNPs reflect true mixed viral populations or sequencing artifacts. We obtained high-quality genomes with minimal variations through these approaches.

### Mutations in F13L gene

Among the 36 sequencing testing conditions assessed, 27 yielded interpretable sequences spanning the F13L gene region, which is associated with tecovirimat resistance. The remaining nine test conditions did not produce sequence data for this gene, all with the metagenomic approach using the Contipharma or the Testsea Ag-RDTs.

## Discussion

Although the number of mpox cases outside of Africa has decreased since 2023, MPXV continues to circulate and affect new countries in Africa. Most recently, an inter-clade recombinant MPXV was detected in England in a traveler recently returning from Asia (27). This event highlights the potential for MPXV to continue to evolve as it circulates in the human population. It is therefore critical to maintain and scale up MPXV genomic surveillance efforts, and to incorporate more data from the most affected areas, even when these regions are difficult to access(28). Metagenomic and tiled amplicon-based Nanopore sequencing of viral genomes is typically performed using nucleic acids extracted directly from clinical specimen (29–31). Previous independent studies on the clinical performance of MPXV Ag-RDTs have shown that they have poor sensitivity(32). However, a recent field evaluation in DRC demonstrated that two of the evaluated RDTs had an improved clinical sensitivity of over 70%(33). This suggests that MPXV Ag-RDTs could be useful in high prevalence contexts as a “rule-in” test, allowing to shorten the time to result and potentially offering new opportunities for genomic surveillance. In this study, we aimed to assess the potential use of Ag-RDTs for genomic surveillance by testing both metagenomic and tiled amplicon-based Nanopore sequencing approaches after DNA extraction from Ag-RDTs.

We demonstrate that MPXV DNA can reliably be extracted from the RDT pads, as Ct values were successfully generated from all positive Ag-RDTs, even after several weeks of storage time at room temperature. As expected, Ct values obtained after pad elution were consistently higher than the initial Ct values measured for each used dilution, indicating that only a fraction of the initial viral genetic material is recovered from the test strip. Moreover, a clear correlation was maintained between initial and post- strip elution Ct values across the three dilution levels, showing that the Ag-RDT strip preserves the relative viral-load gradient despite partial loss during recovery. We also demonstrate the feasibility of sequencing MPXV genomes from MPXV Ag-RDTs stored at RT or +4°C for at least 19 days. This suggests that Ag-RDTs could be transported easily from remote testing sites to centralized diagnostic laboratories for sequencing. Further testing on RDT storage duration and temperature is required to determine acceptable storage conditions prior to DNA extraction and sequencing.

Overall, both sequencing methods achieved good-quality genome coverage, enabling subclade assignment for all samples except one. This sample was analyzed using the metagenomics method and had the highest viral concentration. However, the tiling sequencing approach achieved a higher overall percentage of genome coverage, although very good results have been obtained also with the metagenomic method for some Ag-RDTs. Using the tiled-amplicon protocol, recurrent gaps in identical genomic regions were observed, resulting in a genome coverage never exceeding 94.4%. This pattern was previously observed (informal communication) with similar Ct values, suggesting that the primer design may need to be re-evaluated and/or the PCR protocol optimized to ensure full genome recovery from high Ct values. Although the tiling method achieves better genome coverage, this does not confirm that it is the best sequencing approach in every context. In fact, the metagenomic method is more agnostic and therefore has a better capacity to detect potential viral coinfections(18), as well as new MPXV variants and recombinants, such as the one identified in India and the UK in 2025(7). Although treatment with tecovirimat was not proven to be effective against MPXV in randomized controlled trials and resistance may be of low clinical relevance, we could show successful amplification of the genomic region where mutations associated with tecovirimat resistance have been reported (34, 35).

The overall turnaround times for the metagenomic and tiled-amplicon sequencing approaches were comparable. Although the tiled-amplicon workflow includes a targeted amplification step, the metagenomic protocol used here also involves a PCR step during library preparation. Consequently, both approaches required a similar total processing time from extraction to data generation, with only minor differences in hands-on time. However, the tilling method may allow for faster subclade assignment, as it provides better sequencing depth and likely enables a shorter runtime. Subclade identification has been possible for some samples (not all tested) with an intermediate analysis after 4h sequencing run (data not shown). Faster results can be also achieved by using the recommended ultrafast NextGenPCR^TM^ Thermal Cycler instrument (Molecular Biology Systems).

Our study has some limitations. As this is a preliminary feasibility study, the scope has been limited to the sequencing of the MPXV Ib subclade. Although similar sequencing performance is expected for the other subclades, further investigation is needed to compare and confirm the results for all subclades. Another limitation of this study is that the samples were sequenced 19 days after Ag-RDT testing, having been stored at ambient temperature of less than 25°C under laboratory conditions. In remote areas, storage conditions prior to and during sample shipment could be longer and more extreme. We tested three different Ag-RDTs and two sequencing protocols. Therefore, our results may not be transposable to other MPXV Ag-RDTs and sequencing methods. For NG-Biotech RDT, we had to add an extra step to the sample processing. DNA could not be extracted if the pad was added directly to the lysis buffer. PBS must be added before the lysis buffer. Due to this additional step, a different input volume was used for the nucleic acid extraction (500 µL for Contipharma and Testsea RDTs vs 1000 µL for NG Biotech) but the same elution volume (25 µL) was used for all tests. This variation could have affected the quantity of viral material extracted, therefore impacting genome coverage and sequencing performance. Lastly, we did not use clinical samples. Testing was performed by adding viral culture dilutions to the proprietary buffer of each Ag-RDT. To confirm whether the sequencing performance using clinical specimens would be similar to that observed in our study, testing would need to be performed with cutaneous or mucosal specimens (the validated specimen types for mpox diagnosis) added directly to the Ag-RDT buffer.

Despite these limitations, the sequencing performance was globally of good quality and sufficient to identify the Ib subclade. This study demonstrates that high-quality MPXV genome sequences can be obtained from samples that test positively by Ag-RDTs. These findings address an important need in field settings, demonstrating the feasibility of sequencing from Ag-RDTs, and warrant further evaluation under real-world circumstances.

## Data Availability

All data produced in the present study are available upon reasonable request to the authors

## Contributions

CE, IE, AR and CP conceptualized the project.

KA, VC, FL did data curation and investigation. Formal analysis and methodology development were done by CE, AR and CP.

The original draft was written by CE, AR and CP; review and editing were done by IE.

All authors had full access to all the data in the study and accept responsibility for the decision to submit for publication.

## Acknowledgments

The Geneva Center for Emerging Viral Diseases is a Who Collaborating Centre for Epidemic and Pandemic Diseases. We thank the World Health Organization for providing the diagnostic tests evaluated in this study. We thank the WHO BioHub (Spiez, Switzerland) and the Institut National de Recherche Biomédicale (Kinshasa, Democratic Republic of the Congo) for sharing the MPXV clade Ib isolate.

## References

1. 2022. Epidemiological update: Monkeypox multi-country outbreak. https://www.ecdc.europa.eu/en/news-events/epidemiological-update-monkeypox-multi-country-outbreak. Retrieved 31 March 2025.

2. Lim EY, Whitehorn J, Rivett L. 2023. Monkeypox: a review of the 2022 outbreak. Br Med Bull 145:17–29.

3. 2024. 2022 Mpox Outbreak Global Map | Mpox | Poxvirus | CDC. https://archive.cdc.gov/www_cdc_gov/poxvirus/mpox/response/2022/world-map.html. Retrieved 31 March 2025.

4. O’Toole Á, Neher RA, Ndodo N, Borges V, Gannon B, Gomes JP, Groves N, King DJ, Maloney D, Lemey P, Lewandowski K, Loman N, Myers R, Omah IF, Suchard MA, Worobey M, Chand M, Ihekweazu C, Ulaeto D, Adetifa I, Rambaut A. 2023. APOBEC3 deaminase editing in mpox virus as evidence for sustained human transmission since at least 2016. Science 382:595–600.

5. Akingbola A, Abiodun A, Idahor C, Peters F, Ojo O, Jessica OU, Alao UH, Adewole O, Owolabi A, Chuku J. 2025. Genomic Evolution and Epidemiological Impact of Ongoing Clade Ib MPox Disease: A Narrative Review. Glob Health Epidemiol Genom 2025:8845911.

6. Vakaniaki EH, Kacita C, Kinganda-Lusamaki E, O’Toole Á, Wawina-Bokalanga T, Mukadi-Bamuleka D, Amuri-Aziza A, Malyamungu-Bubala N, Mweshi-Kumbana F, Mutimbwa-Mambo L, Belesi-Siangoli F, Mujula Y, Parker E, Muswamba-Kayembe P-C, Nundu SS, Lushima RS, Makangara-Cigolo J-C, Mulopo-Mukanya N, Pukuta-Simbu E, Akil-Bandali P, Kavunga H, Abdramane O, Brosius I, Bangwen E, Vercauteren K, Sam-Agudu NA, Mills EJ, Tshiani-Mbaya O, Hoff NA, Rimoin AW, Hensley LE, Kindrachuk J, Baxter C, de Oliveira T, Ayouba A, Peeters M, Delaporte E, Ahuka-Mundeke S, Mohr EL, Sullivan NJ, Muyembe-Tamfum J-J, Nachega JB, Rambaut A, Liesenborghs L, Mbala-Kingebeni P. 2024. Sustained human outbreak of a new MPXV clade I lineage in eastern Democratic Republic of the Congo. Nat Med 30:2791–2795.

7. World Health Organization. 2026. Mpox: recombinant virus with genomic elements of clades Ib and IIb – Global situation. https://www.who.int/emergencies/disease-outbreak-news/item/2026-DON595. Retrieved 6 March 2026.

8. Lifting of Mpox as a Public Health Emergency of Continental Security (PHECS) – Africa CDC. https://africacdc.org/news-item/lifting-of-mpox-as-a-public-health-emergency-of-continental-security-phecs/. Retrieved 23 February 2026.

9. Samarasekera U. 2025. WHO ramps up emergency use mpox diagnostics. Lancet Microbe 6:101051.

10. FIND. 2024. DxConnect Test Directory Explorer.

11. WHO. 2023. Target product profiles for tests used for mpox (monkeypox) diagnosis.

12. Ishara-Nshombo E, Somasundaran A, Romero-Ramirez A, Kontogianni K, Mukadi-Bamuleka D, Mukoka-Ntumba M, Muhindo-Milonde E, Mirimo-Nguee H, Parkes J, Hussain Y, Gould S, Williams CT, Wooding D, Nkeramahame J, Watson M, Hardwick HE, Semple MG, Baillie JK, Dunning J, Fletcher TE, Edwards T, Emperador DM, Kavunga-Membo H, Cubas-Atienzar AI, ISARIC 4 and C Investigators. 2025. Diagnostic Accuracy of 3 Mpox Lateral Flow Assays for Antigen Detection, Democratic Republic of the Congo and United Kingdom. Emerg Infect Dis 31:1140–1148.

13. Adea K, Escadafal C, Emperador DM, Agogo E, O’Driscoll M, Mbala-Kingebeni P, Kaiser L, Bekliz M, Eckerle I. 2026. Comparative analytical sensitivity evaluation of five mpox point-of-care assays for mpox clades Ia, Ib, IIa and IIb. medRxiv 2026.01.17.26344231.

14. Smith TG, Gigante CM, Wynn NT, Matheny A, Davidson W, Yang Y, Condori RE, O’Connell K, Kovar L, Williams TL, Yu YC, Petersen BW, Baird N, Lowe D, Li Y, Satheshkumar PS, Hutson CL. 2023. Tecovirimat Resistance in Mpox Patients, United States, 2022-2023. Emerg Infect Dis 29:2426–2432.

15. Postal J, Guivel-Benhassine F, Porrot F, Grassin Q, Crook JM, Vernuccio R, Caro V, Vanhomwegen J, Guardado-Calvo P, Simon-Lorière E, Dacheux L, Manuguerra J-C, Schwartz O. 2025. Antiviral activity of tecovirimat against monkeypox virus clades 1a, 1b, 2a, and 2b. The Lancet Infectious Diseases 25:e126–e127.

16. Lagathu G, Grolhier C, Besombes J, Maillard A, Comacle P, Pronier C, Thibault V. 2023. Using Discarded Facial Tissues to Monitor and Diagnose Viral Respiratory Infections. Emerg Infect Dis 29:511–518.

17. Rector A, Bloemen M, Van Ranst M, Wollants E. 2023. Used paper tissues for pathogen identification in acute respiratory infection. J Med Virol 95:e29127.

18. Pérez-Rodríguez F-J, Laubscher F, Chudzinski V, Kaiser L, Cordey S. 2023. Direct Dengue Virus Genome Sequencing from Antigen NS1 Rapid Diagnostic Tests: A Proof-of-Concept with the Standard Q Dengue Duo Assay. Viruses 15:2167.

19. Paull JS, Petros BA, Brock-Fisher TM, Jalbert SA, Selser VM, Messer KS, Dobbins ST, DeRuff KC, Deng D, Springer M, Sabeti PC. 2024. Optimisation and evaluation of viral genomic sequencing of SARS-CoV-2 rapid diagnostic tests: a laboratory and cohort-based study. Lancet Microbe 5:e468–e477.

20. Moso MA, Taiaroa G, Steinig E, Zhanduisenov M, Butel-Simoes G, Savic I, Taouk ML, Chea S, Moselen J, O’Keefe J, Prestedge J, Pollock GL, Khan M, Soloczynskyj K, Fernando J, Martin GE, Caly L, Barr IG, Tran T, Druce J, Lim CK, Williamson DA. 2024. Non-SARS-CoV-2 respiratory viral detection and whole genome sequencing from COVID-19 rapid antigen test devices: a laboratory evaluation study. Lancet Microbe 5:e317–e325.

21. Nazario-Toole A, Nguyen HM, Xia H, Frankel DN, Kieffer JW, Gibbons TF. 2022. Sequencing SARS-CoV-2 from antigen tests. PLoS One 17:e0263794.

22. Rector A, Bloemen M, Schiettekatte G, Maes P, Van Ranst M, Wollants E. 2023. Sequencing directly from antigen-detection rapid diagnostic tests in Belgium, 2022: a gamechanger in genomic surveillance? Euro Surveill 28:2200618.

23. Scaramozzino N, Ferrier-Rembert A, Favier A-L, Rothlisberger C, Richard S, Crance J-M, Meyer H, Garin D. 2007. Real-time PCR to identify variola virus or other human pathogenic orthopox viruses. Clin Chem 53:606–613.

24. protocol_for_the_detection_of_monkeypox_by_rt.pdf.

25. Welkers M, Jonges M, Van Den Ouden A. 2022. Monkeypox virus whole genome sequencing using combination of NextGenPCR and Oxford Nanopore v1 10.17504/protocols.io.n2bvj6155lk5/v1.

26. Treutiger C-J, Filén F, Rehn M, Aarum J, Jacks A, Gisslén M, Sturegård E, Karlberg ML, Karlsson Lindsjö O, Sondén K. 2024. First case of mpox with monkeypox virus clade Ib outside Africa in a returning traveller, Sweden, August 2024: public health measures. Euro Surveill 29:2400740.

27. Steven T. Pullan, Isobel Everall, Rebecca Doherty, Lucy Crossman, Emma Wise, Hassan Hartman, Natalie Groves, Dominic Worku, Catherine Houlihan, Tommy Rampling, Claire Gordon, Meera Chand, Andrew Rambaut. 2025. Inter-Clade Recombinant Mpox Virus Detected in England in a Traveller Recently Returned from Asia.

28. Kinganda-Lusamaki E, Amuri-Aziza A, Fernandez-Nuñez N, Makangara-Cigolo J-C, Pratt C, Vakaniaki EH, Hoff NA, Luakanda-Ndelemo G, Akil-Bandali P, Nundu SS, Mulopo-Mukanya N, Ngimba M, Modadra-Madakpa B, Diavita R, Paku-Tshambu P, Pukuta-Simbu E, Merritt S, O’Toole Á, Low N, Nkuba-Ndaye A, Kavunga-Membo H, Shongo Lushima R, Liesenborghs L, Wawina-Bokalanga T, Vercauteren K, Mukadi-Bamuleka D, Subissi L, Muyembe-Tamfum J-J, Kindrachuk J, Ayouba A, Rambaut A, Delaporte E, Tessema S, D’Ortenzio E, Rimoin AW, Hensley LE, Mbala-Kingebeni P, Peeters M, Ahuka-Mundeke S. 2025. Clade I mpox virus genomic diversity in the Democratic Republic of the Congo, 2018-2024: Predominance of zoonotic transmission. Cell 188:4-14.e6.

29. Kwasiborski A, Hourdel V, Balière C, Hoinard D, Grassin Q, Feher M, De La Porte Des Vaux C, Cresta M, Vanhomwegen J, Manuguerra J-C, Batéjat C, Caro V. 2024. Direct metagenomic and amplicon-based Nanopore sequencing of French human monkeypox from clinical specimen. Microbiol Resour Announc 13:e00811–23.

30. Vandenbogaert M, Kwasiborski A, Gonofio E, Descorps-Declère S, Selekon B, Nkili Meyong AA, Ouilibona RS, Gessain A, Manuguerra J-C, Caro V, Nakoune E, Berthet N. 2022. Nanopore sequencing of a monkeypox virus strain isolated from a pustular lesion in the Central African Republic. Sci Rep 12:10768.

31. Schuele L, Boter M, Nieuwenhuijse DF, Götz H, Fanoy E, de Vries H, Vieyra B, Bavalia R, Hoornenborg E, Molenkamp R, Jonges M, van den Ouden A, Simões M, van den Lubben M, Koopmans M, Welkers MRA, Oude Munnink BB. 2024. Circulation, viral diversity and genomic rearrangement in mpox virus in the Netherlands during the 2022 outbreak and beyond. J Med Virol 96:e29397.

32. Ishara-Nshombo E, Somasundaran A, Romero-Ramirez A, Kontogianni K, Mukadi-Bamuleka D, Mukoka-Ntumba M, Muhindo-Milonde E, Mirimo-Nguee H, Parkes J, Hussain Y, Gould S, Williams CT, Wooding D, Nkeramahame J, Watson M, Hardwick HE, Semple MG, Baillie JK, Dunning J, ISARIC 4, C Investigators, Fletcher TE, Edwards T, Emperador DM, Kavunga-Membo H, Cubas-Atienzar AI. 2025. Diagnostic Accuracy of 3 Mpox Lateral Flow Assays for Antigen Detection, Democratic Republic of the Congo and United Kingdom. Emerg Infect Dis 31.

33. Bakamutumaho B, Mwenebitu DL, Muttamba W, Ampeire I, Bosa HK, Bahizire E, Bisimwa BC, Obuku A, Wayengera M, Sandeman A, Holden MTG, Katoto PDMC, Kirenga B, Williamson DA, Sabiiti W. 2025. Field evaluation of a rapid antigen test for mpox in the Democratic Republic of the Congo and Uganda: a multicentre, prospective, diagnostic accuracy study. Lancet Infect Dis S1473–3099(25)00600–0.

34. The PALM007 Writing Group. 2025. Tecovirimat for Clade I MPXV Infection in the Democratic Republic of Congo. N Engl J Med 392:1484–1496.

35. Zucker J, Fischer WA, Zheng L, McCarthy C, Saha PT, Javan AC, Greninger A, Hamill MM, Leslie K, Brooks KM, Berardi J, Smith D, Hosey L, Aldrovandi G, Ferbas K, Day C, Bender Ignacio RA, Bolan R, Glesby MJ, Landovitz RJ, Luetkemeyer AF, Sierra Madero J, Gandhi RT, Nachman S, Eron J, Currier JS, Wilkin T. 2026. Tecovirimat for the Treatment of Mpox. N Engl J Med 394:884–895.

